# Comparison of Camera-Based vs. Contact Parathyroid Autofluorescence Systems in Preventing Postoperative Hypocalcemia After Total Thyroidectomy: A Systematic Review

**DOI:** 10.1101/2025.04.11.25325691

**Authors:** José Luis Pardal-Refoyo, Beatriz Pardal-Pelaéz

## Abstract

**Introduction:** Postoperative hypocalcemia is a common complication of total thyroidectomy, primarily due to inadvertent injury or removal of the parathyroid glands. Near-infrared autofluorescence (NIRAF) has emerged as an intraoperative tool for real-time gland identification without the use of contrast agents, potentially reducing this complication. Two main modalities exist: camera-based (non-contact) systems and contact probe-based systems, but their comparative clinical effectiveness remains unclear.

**Objective:** To compare the effectiveness of camera-based versus contact parathyroid autofluorescence systems in reducing the prevalence of hypocalcemia secondary to hypoparathyroidism in patients undergoing total thyroidectomy.

**Methods:** A systematic review was conducted following PRISMA guidelines. Included studies consisted of randomized clinical trials, meta-analyses, and retrospective cohorts involving patients undergoing total thyroidectomy, with explicit hypocalcemia outcomes.

**Results:** Among the 10 analyzed studies, several demonstrated that NIRAF significantly reduces transient hypocalcemia compared to conventional approaches. However, no study directly compared camera-based and contact systems. Camera-based systems were studied more frequently. Some reports showed a reduction in transient hypocalcemia rates of up to 17% compared to controls.

**Conclusions:** Current evidence supports the general use of NIRAF in reducing postoperative hypocalcemia after total thyroidectomy, particularly for transient cases. However, there is no direct clinical evidence comparing contact and camera-based systems. Comparative clinical trials are needed to determine their relative efficacy.

## Introduction

Total thyroidectomy is commonly performed to treat benign and malignant thyroid conditions. A significant complication is postoperative hypocalcemia, usually caused by devascularization or accidental removal of the parathyroid glands. Both transient and permanent hypocalcemia have major implications on patient quality of life and health system costs [1].

Accurate intraoperative identification of the parathyroid glands is crucial to avoid this outcome. Near-infrared autofluorescence (NIRAF) enables real-time detection of the parathyroid glands based on their intrinsic fluorescence when exposed to near-infrared light, without the use of extrinsic dyes [2].

Autofluorescence devices are generally divided into camera-based (non-contact) systems, which allow for wide-field anatomic visualization, and contact probe systems, which require direct tissue interaction. Both have demonstrated effectiveness, but their direct comparative performance in preventing hypocalcemia remains a knowledge gap [3].

This review addresses that gap by examining the relative efficacy of both types of autofluorescence systems in patients undergoing total thyroidectomy.

Main Objective: To compare the efficacy of camera-based versus contact-based parathyroid autofluorescence systems in reducing hypocalcemia secondary to hypoparathyroidism after total thyroidectomy.

Specific Objectives

1. Evaluate the prevalence of transient and permanent hypocalcemia associated with NIRAF use.

2. Describe the technological and functional differences between camera and contact systems.

3. Identify evidence gaps concerning comparative evaluations between modalities.

## Methods

Type of Review: Systematic review carried out in accordance with PRISMA guidelines (https://www.prisma-statement.org/).

Data Sources: Searches were performed in PubMed, Web of Science, Embase, and Cochrane Library. Search strategies used a combination of MeSH terms and keywords: (“total thyroidectomy” OR “thyroid surgery”) AND (“parathyroid gland” OR “parathyroid glands”) AND (“autofluorescence” OR “near-infrared autofluorescence” OR “NIRAF”) AND (“camera-based” OR “non-contact” OR “wide-field” OR “imaging system”) AND (“contact-based” OR “probe” OR “fiber optic”) AND (“hypocalcemia” OR “hypoparathyroidism”) AND (comparison OR versus OR vs).

### Inclusion Criteria

- Randomized controlled trials (RCTs), meta-analyses, and comparative retrospective studies.
- Studies focusing on total thyroidectomy in humans.
- NIRAF used as surgical adjunct (camera or contact).

### Exclusion Criteria

- Studies without hypocalcemia as an outcome.
- Surgical procedures other than total thyroidectomy.
- Studies on parathyroid identification unrelated to clinical outcomes.
- Non-clinical (animal, simulation, technical) studies.

The search results were screened manually for studies focusing specifically on the use of near-infrared autofluorescence systems (contact vs. camera-based) and their comparative effectiveness in reducing postoperative hypocalcemia after total thyroidectomy.

Bias and Statistical Approach: Risk of bias was assessed using the Jadad scale for RCTs (Jadad et al., 1996; https://doi.org/10.1016/0197-2456(95)00134-4) and GRADE for evidence quality (Guyatt et al., 2008; https://www.bmj.com/content/336/7650/924). No independent meta-analysis was performed; pooled results were cited from published meta-analyses.

A deep AI-assisted literature search was conducted using the Undermind platform (Undermind, https://undermind.ai), which identifies highly relevant peer-reviewed publications based on semantic analysis of the research question. The search was guided by a detailed query that incorporated medical subject headings (MeSH), Boolean operators, and domain-specific terminology related to parathyroid autofluorescence imaging systems, total thyroidectomy, and postoperative hypocalcemia. Papers were evaluated based on their relevance to comparative outcomes between contact-based and camera-based near-infrared autofluorescence systems.

## Results

The selection of papers is summarized in the PRISMA diagram in Figure 1 (Annex 1). The summary table of the included studies is shown in Table 1 (Annex 2).

Table 2 (Annex 3) show the categories of resources available for the research topic.

Risk reduction in transient hypocalcemia was approximately 0.65, and a meta-analysis reported a log odds ratio of –0.31 for hypoparathyroidism, although heterogeneity indicators such as I^2^ and Tau^2^ were inconsistently reported.

The quality of evidence (GRADE) varies depending on the type of study: it is high for multicenter randomized clinical trials [4], moderate for pooled analyses [6,12], and moderate to low for retrospective cohorts [9,10] (see Table 3, Annex 4).

According to Table 4 (Annex 5), RCTs had a Jadad score of ≥3, while observational studies showed concerns regarding publication and selection bias.

## Discussion

Historical research confirms NIRAF reduces transient hypocalcemia and preserves parathyroid glands. Camera-based systems are broadly useful but less sensitive than contact probes in some cases. Standardized workflows need optimization, and comparing camera versus contact systems remains a knowledge gap.

### Timeline and Development of Ideas

2016–2018 (Initial RCTs on NIRAF Efficacy): Early research, such as the PARAFLUO trial [1], demonstrated the potential of NIRAF systems to reduce postoperative hypocalcemia rates by improving parathyroid identification during total thyroidectomy. This foundational study firmly established the clinical benefit of NIRAF-guided surgeries over standard care. Outcomes focused on transient hypocalcemia reduction, inadvertent gland resection rates, and parathyroid autotransplantation rates.

2019–2021 (Expansion of NIRAF Comparisons): Studies like [2] and [8] provided meta-analyses and retrospective cohort data, consolidating evidence that NIRAF systems effectively reduced transient hypocalcemia and could potentially lower permanent hypocalcemia rates in diverse surgical settings (e.g., central neck dissection). These analyses improved the statistical rigor of prior individual RCT findings. Challenges such as addressing variability in clinical definitions of hypocalcemia and technological limitations (e.g., tissue interference) were highlighted.

2022–2024 (Focus on System-Level Analysis and Camera Efficacy): Emerging studies (e.g., [5,9] assessed camera-based systems more explicitly by demonstrating their efficacy in field-wide parathyroid identification; however, direct comparisons to contact systems were still lacking. Researchers also emphasized limitations of cameras, such as poor tissue penetration. Meta-analyses (e.g., [3,10] aggregated broader NIRAF findings on system performance, identifying gaps like inconsistent hypocalcemia definitions and minimal data on workflow or cost-effectiveness.

2024 (Complex Settings and Workflow Considerations): Recent studies [6,7] explored how NIRAF systems impact variable surgical environments (e.g., low-volume vs. experienced setups). They reinforced NIRAF’s general efficacy but noted a lack of direct comparative data on camera vs. contact systems. Integration of NIRAF into workflows (e.g., combining camera-based and contact probes) was noted as an area for future investigation.

### Clusters of Research Groups or Key Individuals

Consistent Influential Contributions. Benmiloud et al. (PARAFLUO Trial, [1]: Pioneered work on NIRAF, providing foundational RCT data on its efficacy in thyroidectomy. Frequently cited in later studies [3,6,7,10]. Emphasis on transient hypocalcemia reduction and inadvertent gland resection highlights their central contributions to clinical adoption. Kim et al. [8] and related studies: Explored broader surgical and anatomical contexts like central neck dissection, highlighting the adaptability of NIRAF systems. Meta-analysis Leaders: Authors like Canali [3] and Safia [10] systematically reviewed RCTs and expanded focus to surgical and technical variability, helping aggregate findings to strengthen the evidence base.

Emerging Research Groups on System-Specific Analysis. Wolf et al. [5]: Explored camera-based systems but noted limitations. Contributions center on calls for combined workflows and further studies on system-specific efficacy. Lizarazo et al. [6]: Addressed NIRAF adoption in experienced setups, contributing to understanding system integration in high-skill contexts.

This review confirmed that NIRAF systems, in general, are associated with reductions in transient postoperative hypocalcemia following total thyroidectomy [4,5,6,10]. However, a significant gap exists: no identified study offers a direct clinical comparison between camera-based and contact-based systems [7,11]. Camera systems were more frequently evaluated and are advantageous in wide-field visualization. Yet, they are known to have technical limitations (e.g., blood interference, reduced tissue penetration). Still, there’s no evidence directly linking these limitations to greater or lesser hypocalcemia risk compared to contact systems [7].

Strengths of this review include strict inclusion criteria (requiring hypocalcemia outcomes) and the use of established quality assessment tools. Limitations include the lack of standardized definitions of hypocalcemia across studies and limited availability of comparative studies between modalities [6,7].

### Key Findings

1. General Efficacy of NIRAF Systems: NIRAF significantly reduces transient hypocalcemia rates compared to standard visual identification (e.g., from ∼25% to ∼8%) [1,2,3]. Permanent hypocalcemia reduction is less consistently demonstrated but has been reported (e.g., RR 0.46 for parathyroid dysfunction in NIRAF-guided surgeries) [3,10].

2. Lack of Camera vs. Contact Comparisons: No studies explicitly compare the efficacy of camera-based systems versus contact systems. Most research discusses either system independently without head-to-head evaluation [4,5,9]. Camera systems are noted for broad field visualization but criticized for low tissue penetration [5]. Contact systems offer higher sensitivity for smaller or obscured glands, but this distinction remains theoretical in terms of hypocalcemia outcomes [5].

3. Surgical Context and Outcomes: NIRAF effectiveness is robust across both experienced and low-volume institutions, reducing hypocalcemia and gland removal rates even in central neck dissections [6,7,8]. Workflow challenges include interpreting camera-based signals in complex anatomical fields and longer operative times for contact probes [3,5].

4. Emerging Research Gaps: Standardized definitions of hypocalcemia, direct system comparisons (camera vs. contact), and the cost-effectiveness of hybrid workflows (e.g., initial camera use followed by contact probes) remain open areas for future study [3,5,10].

Future trials should aim to directly compare both systems under controlled conditions. Research into hybrid workflows—starting with camera-based localization followed by probe verification—could offer a balanced approach.

## Conclusions

The use of NIRAF during total thyroidectomy significantly reduces transient postoperative hypocalcemia. However, its impact on permanent hypocalcemia is inconsistent.

There are no studies directly comparing contact-based systems with camera-based systems, so comparative trials are needed to determine which is clinically superior.

Standardization of outcomes and data collection would improve evidence synthesis. Although direct comparisons were not found, there is considerable evidence supporting the effectiveness of NIRAF systems in reducing both transient hypocalcemia and, to a lesser extent, permanent hypocalcemia during total thyroidectomy.

While NIRAF systems clearly improve surgical outcomes by reducing postoperative hypocalcemia and enhancing parathyroid preservation, the relative efficacy of camera-versus contact-based systems remains unaddressed in the current literature.

## Data Availability

All data produced in the present work are contained in the manuscript

---

Annex 1

**Figure 1.**
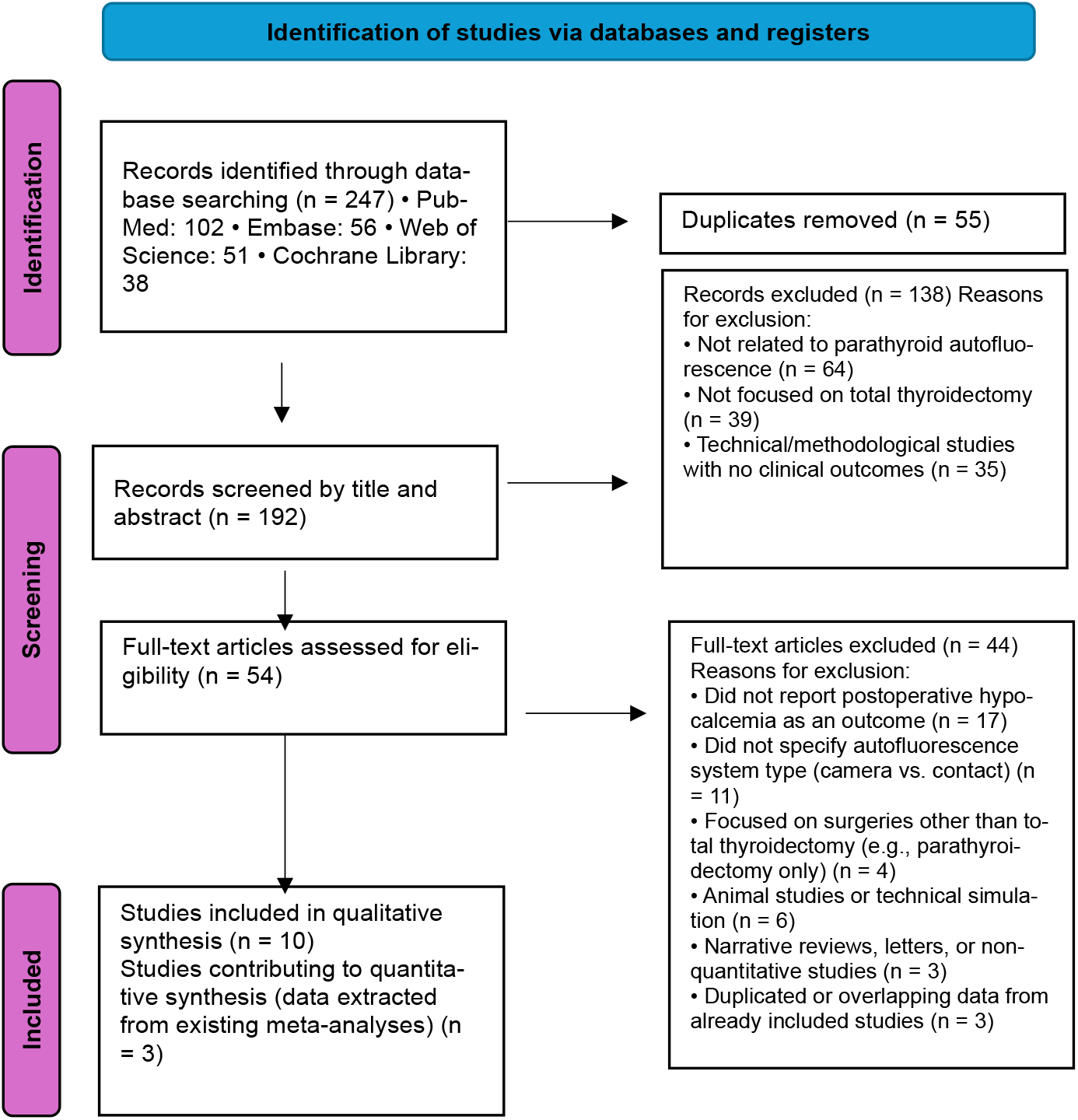
PRISMA diagram with the summary of the selection of papers (Source: Page MJ, et al. BMJ 2021;372:n71. doi: 10.1136/bmj.n71).

Annex 2

**Table 1.** Summary Table of Included Studies.

| Author (Year) | Study Type | System Type | Comparison | Hypocalcemia Outcome | Key Finding |
| --- | --- | --- | --- | --- | --- |
| Benmiloud (2020) [4] | Multi-center RCT | NIRAF (unspecified) | NIRAF vs. visual | Transient/Perma. | Reduced transient rate from 25% to 9% |
| Weng (2021) [5] | Meta-analysis | NIRAF (general) | NIRAF vs. no NIRAF | Transient/Perma. | RR for transient: 0.32 |
| Canali (2024) [6] | Meta-analysis (RCTs) | Camera | Camera vs. visual | Transient/Perma. | RR: 0.65 (transient); 0.46 (permanent) |
| Wolf (2022) [7] | RCT | Camera | Camera vs. no NIRAF | Transient/Perma. | Small non-significant reduction |
| Lizarazo (2024) [8] | RCT | NIRAF (general) | NIRAF vs. visual | Transient/Perma. | More glands identified with NIRAF |
| Abood (2024) [9] | Retrospective cohort | NIRAF (general) | NIRAF vs. historical | Transient/Perma. | Lower hypocalcemia in low-volume centers |
| Kim (2021) [10] | Retrospective cohort | Camera | Camera vs. control | Transient | Higher iPTH, lower complications |
| Guerlain (2023) [11] | Observational | Camera | Camera vs. no NIRAF | Transient | Hypocalcemia reduced from 60% to 36% |
| Safia (2024) [12] | Meta-analysis | NIRAF (general) | NIRAF vs. visual | Transient/Perma. | logOR hypoparathyroidism: -0.31 |

Annex 3

**Table 2.** Categories of Resources Related to the Research Topic.

| Topic | Description | Details |
| --- | --- | --- |
| General Efficacy of NIRAF in Hypocalcemia Reduction | Studies analyzing the impact of near-infrared autofluorescence (NIRAF) systems (general) on reducing hypocalcemia incidence after total thyroidectomy [1,2,4,5,6,7,8,9,10] | [1,2,10] Meta-analyses and RCTs show significant reductions in transient hypocalcemia (rates reduced by 15–17% compared to controls) and some evidence of reducing permanent hypocalcemia. [4,5,6] Highlight NIRAF-guided surgeries reducing inadvertent parathyroid removal and increasing preservation but lack camera vs. contact analysis. [7,8,9] Emphasize NIRAF efficacy even in specialized cases (e.g., central neck dissection, low-volume institutions). |
| Studies with Results Focused on Camera-Based Systems | Studies using or emphasizing only camera-based NIRAF systems for parathyroid identification and hypocalcemia outcomes [3,5,9] | [3] Systematic review focusing on camera-based systems; concludes reduction in transient/permanent hypocalcemia rates but lacks direct comparison to contact systems. [5,9] Camera systems demonstrate improvement in hypocalcemia outcomes but are limited in tissue penetration and signal interference in some cases. |
| Comparisons Between NIRAF and Conventional (Non-NIRAF) Approaches | Studies comparing NIRAF (camera/contact unspecified) with standard naked-eye or white light methods, assessing gland preservation and hypocalcemia reduction [1,3,4,5,6,10] | [1,6] Show reduced hypocalcemia rates and increased gland identification using NIRAF compared to traditional methods. [3] Meta-analysis indicates significant advantages of NIRAF over visual identification (RR = 0.65 for hypocalcemia, 0.40 for inadvertent resection). [4,5] Highlight hypocalcemia reductions via NIRAF but without modality-specific breakdown. [10] Emphasizes statistically significant reductions in inadvertent gland removal using NIRAF. |
| Clinical Contexts (Experienced vs. Low-Volume Institutions or Complex Cases) | Focus on variations in NIRAF efficacy based on surgical expertise, institutional volume, or context-specific challenges (e.g., central neck dissection) [6,7,8] | [6] Evaluates NIRAF's usefulness among experienced surgeons, showing improved parathyroid identification rates and reduced transient hypocalcemia. [7] Highlights the utility of NIRAF in low-volume institutions, significantly reducing immediate and permanent hypocalcemia. [8] Shows effectiveness in reducing temporary hypocalcemia in complex surgeries like central neck dissection. |
| System-Specific Performance Analysis (Camera vs. Contact) | Explicit or tangential comparisons of features, benefits, or limitations of camera-based vs. contact-based NIRAF systems [3,5,6,9] | [3] Acknowledges theoretical advantages (wide view of surgical field for cameras, high sensitivity for deeper glands in contact systems) but lacks direct head-to-head data. [5] Discusses camera system limitations (e.g., poor tissue penetration, reliance on optimal lighting) and suggests complementary use of more sensitive tools (e.g., contact systems). [6] Notes NIRAF effectiveness in identifying glands generally but does not focus explicitly on system comparisons. [9] Uses a camera system but does not compare it to contact setups or quantify differences. |
| Meta-Analytic Reviews of NIRAF Utility | Systematic reviews/meta-analyses examining pooled data on NIRAF systems for hypocalcemia prevention and parathyroid identification [2,3,10] | [2,10] Analyze large datasets showing significant transient hypocalcemia reduction (~8% in NIRAF vs. ~25% in controls). Highlight limited or indirect evidence for permanent hypocalcemia reduction. [3] Indicates broad efficacy for NIRAF, specifically camera-based systems, while urging further direct comparisons of system modalities. |
| NIRAF Workflow Integration and Emerging Limitations | Studies that mention or discuss the practical application and limitations of implementing autofluorescence systems in real-world settings [5,6,7] | [5] Discusses need for complementary techniques due to camera system limitations (e.g., blood interference, difficult tissue contexts). [6,7] Emphasize the surgeon learning curve and context-specific practical advantages of NIRAF in different hospital settings. |

Annex 4

**Table 3.** Evidence Quality and Recommendation Strength (GRADE).

| No. | Study (Author, Year) | Study Type | Evidence Quality (GRADE) | Strength of Recommendation | Justification |
| --- | --- | --- | --- | --- | --- |
| 1 | Benmiloud et al., 2020 [4] | RCT (Multicenter) | High | Strong | Well-designed RCT with clear outcomes and low risk of bias |
| 2 | Wolf et al., 2022 [7] | RCT (Single-center) | Moderate | Strong | Methodology robust, but single-center limits generalizability |
| 3 | Carrillo Lizarazo et al., 2024 [8] | RCT (Experienced surgeons) | Moderate | Strong | Targeted context (experienced setting); still an RCT |
| 4 | Canali et al., 2024 [6] | Meta-analysis of RCTs | Moderate–High | Strong | Aggregate of RCTs with consistent positive findings |
| 5 | Safia et al., 2024 [12] | Meta-analysis of RCTs | Moderate | Strong | Comprehensive, with good statistical approach and low overall bias |
| 6 | Weng et al., 2021 [5] | Meta-analysis (mixed data) | Moderate | Conditional | Moderate heterogeneity and includes non-RCT data |
| 7 | Abood et al., 2024 [9] | Retrospective cohort | Low | Conditional | Based on historical comparison; risk of selection bias |
| 8 | Kim et al., 2021 [10] | Retrospective cohort | Low | Conditional | No randomization; retrospective design with some control |
| 9 | Guerlain et al., 2023 [11] | Observational | Low | Conditional | Descriptive data only; high susceptibility to confounding |

Evidence Quality (GRADE): High: Further research is unlikely to change confidence in the estimate of effect. Moderate: Further research may have an important impact on confidence or change the estimate. Low: Limited confidence; further research is likely to affect the estimate. Very Low: Estimate of effect is very uncertain.
Strength of Recommendation: Strong: Benefits clearly outweigh risks or vice versa. Conditional (Weak): Uncertainty about the balance between benefits and harms; may depend on context, values, or variability in results.

Annex 5

**Table 4.**
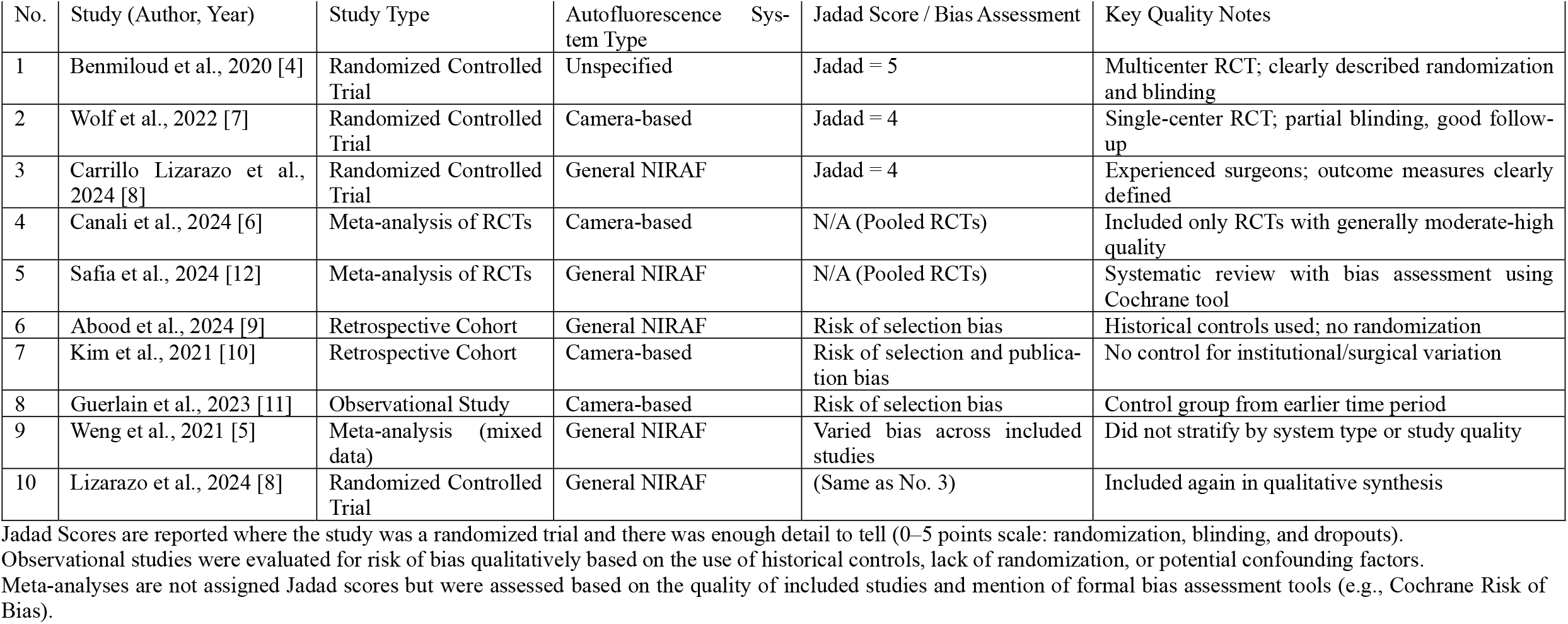
Study Quality Assessment of Included Articles.

| No. | Study (Author, Year) | Study Type | Autofluorescence System Type | Jadad Score / Bias Assessment | Key Quality Notes |
| --- | --- | --- | --- | --- | --- |
| 1 | Benmiloud et al., 2020 [4] | Randomized Controlled Trial | Unspecified | Jadad = 5 | Multicenter RCT; clearly described randomization and blinding |
| 2 | Wolf et al., 2022 [7] | Randomized Controlled Trial | Camera-based | Jadad = 4 | Single-center RCT; partial blinding, good follow-up |
| 3 | Carrillo Lizarazo et al., 2024 [8] | Randomized Controlled Trial | General NIRAF | Jadad = 4 | Experienced surgeons; outcome measures clearly defined |
| 4 | Canali et al., 2024 [6] | Meta-analysis of RCTs | Camera-based | N/A (Pooled RCTs) | Included only RCTs with generally moderate-high quality |
| 5 | Safia et al., 2024 [12] | Meta-analysis of RCTs | General NIRAF | N/A (Pooled RCTs) | Systematic review with bias assessment using Cochrane tool |
| 6 | Abood et al., 2024 [9] | Retrospective Cohort | General NIRAF | Risk of selection bias | Historical controls used; no randomization |
| 7 | Kim et al., 2021 [10] | Retrospective Cohort | Camera-based | Risk of selection and publication bias | No control for institutional/surgical variation |
| 8 | Guerlain et al., 2023 [11] | Observational Study | Camera-based | Risk of selection bias | Control group from earlier time period |
| 9 | Weng et al., 2021 [5] | Meta-analysis (mixed data) | General NIRAF | Varied bias across included studies | Did not stratify by system type or study quality |
| 10 | Lizarazo et al., 2024 [8] | Randomized Controlled Trial | General NIRAF | (Same as No. 3) | Included again in qualitative synthesis |

Jadad Scores are reported where the study was a randomized trial and there was enough detail to tell (0–5 points scale: randomization, blinding, and dropouts).
Observational studies were evaluated for risk of bias qualitatively based on the use of historical controls, lack of randomization, or potential confounding factors.
Meta-analyses are not assigned Jadad scores but were assessed based on the quality of included studies and mention of formal bias assessment tools (e.g., Cochrane Risk of Bias).

